# Risk Factors Associated with Falls in Older Adults with Dementia and Alzheimer’s Diseases among Older Adults in the United States

**DOI:** 10.1101/2023.01.10.23284411

**Authors:** Tayebeh Baniasadi

## Abstract

**Objective:** This study aimed to examine the risk of fall, fall injury, and fall numbers among older adults with dementia and Alzheimer diseases. Additionally, this study explored the relationship of falls by medication use of neurodegenerative diseases.

**Methods:** The survey data from the years 2020 of Health and Retirement Study Consumption and Activities Mail Survey (HRS CAMS) was used. The HRS CAMS includes information of demographic characteristics, fall information, and medical background including dementia and Alzheimer diseases, and medication record. A regression model was used to test whether neurodegenerative diseases and medications was associated with fall risk factors.

**Results:** The sample (n = 8782) was predominately female (54.7%) and white (60.7%) with a mean age of 70.4 years. When controlled for covariates, the findings show 10 percent higher risk of fall for elderly people with dementia and Alzheimer. People with dementia were 9% more likely to have higher risk of injury by fall and those with dementia and Alzheimer’s had more than 7 times of higher chance of a higher number of falls. Using Alzheimer’s prescription was associated with lower 90% lower risk of fall than controls.

**Conclusions:** Dementia and Alzheimer diseases are significant risk factor for falls in older adults. This study suggests that older adults with neurodegenerative diseases have higher risk, and needs more regular medical checkups to decrease the risk of fall.

## Introduction

Falls among the elderly are one of the most frequent reasons for morbidity and mortality, which have important clinical, socioeconomic, and public health problems^1-3^. A wide variety of studies have examined the causes and risk factors of falls in the elderly^4^. One of the main risk factors is cognitive impairment, older with cognitive impairment are more likely to have fall risk^5-6^. However, to my best of knowledge, few research examined whether falls is related to prescriptions for memory problem among older adults.

It is generally believed that walking is an automatic process requiring little or no higher cognitive input^7^. Falls are twice as likely to occur in elderly with dementia and Alzheimer Diseases (AD) as in older adults without those, most likely because of motor impairment, attentional deficits, psychotropic medication, or behavioral symptoms^8-11^. Other risk factors are the lack of a walking aid, functional impairment, recurrent falls, and living in long-term residence^12-15^. Therefore, walking under normal circumstances may require attention and executive function, and cognitive changes could increase the risk of falling^16^.

Use of medications for memory problems are highly criticized due to inappropriate prescription, insufficient risk-benefit balance, and lack of regular evaluation ^17,24^. Medications for memory problems are known to have adverse effects, such as falls^18, 23^. Previous research shows use of memory problem medications has been associated with increased risk of falls in geriatric care settings^19^.

The objective of this study is to identify the risk of fall, fall injury, and fall numbers among older adults with dementia and AD. In addition, this study assessed the relationship of falls by medication use of neurodegenerative diseases. This is expected that having dementia and AD is associated with higher fall risk, injuries, and medication use can be a protective factor to reduce fall risk and injuries.

### Method

Health and Retirement Study (HRS) data was used for this study, which is a longitudinal survey of households with at least one adult over the age of 50 that is nationally representative. The survey is funded by the National Institute on Aging (NIA U01AG009740) and has been directed by the University of Michigan since 1992. Every two years, the survey components are conducted (either face-to-face or by telephone), resulting in a sample of approximately 20,000 respondents with an overall response rate of more than 80%. Initially, the study aimed to examine the effects of health, family, and economic variables during the transition to retirement. Demographic information, healthcare use, work status, family structure, mental and physical health, and finances are all included in the survey. Deidentified data are available for download from the HRS website (https://hrs.isr.umich.edu/data-products). In the present study, we used data from the biennial core survey in 2020, and we included individuals over 65.

### Measures

#### Outcomes

The outcomes of this study were falls, the number of falls and fall injuries. First, falls were defined by the question “Have you fallen down in the last two years? “ and response options were “yes,” “no,” “don’t know,” or “refused to answer.” Those responding “yes” were classified as a faller whereas those answering “no” were classified as non-fallers. For those stating they had fallen in the past 2 years, a follow-up question was asked about the number of falls and injuries. For the number of falls, they were asked “ How many times have you fallen”, and for fall injuries, they were asked, “In that fall/In any of these falls, did you injure yourself seriously enough to need medical treatment?”. In all of the falls-related questions, “don’t know” and “refused to answer” were excluded.

### Independent Variables

This study has four main independent variables. First, is Alzheimer’s Disease (AD) diagnosis. The respondents were asked, “Since we last asked you has a doctor told you that you have Alzheimer’s Disease?”. Those who answered “yes” were coded for ADs patients. In the follow-up question, they were asked “ Are you taking any medication prescribed by a doctor to help with your Alzheimer’s Disease?”. Patients who answered “yes” to this question were classified as those who used an AD prescription. For dementia, they were asked, “Since we last asked you, has a doctor told you that you have dementia, senility or any other serious memory impairment?”. Patients who answered “yes” were coded as having dementia. Finally, the survey asked a more general question for the memory problem, which was “Are you taking any medication prescribed by a doctor to help with your [Alzheimer’s disease/dementia, senility or memory impairment/memory problems]?”. People who answered “yes” to the question were also coded using a memory prescription.

### Covariates

Potential confounding variables related to falls and fall-related injuries included demographic variables and important health-related factors. Demographic variables are age, sex, race, and years of education. Health-related factors confounders were self-reported eyesight, smoking status self-rated health, and frequency of vigorous physical activity.

## Results

Among 8782 participants in this study, nearly 19.5 percent of them reported fall, and 6 percent of had injury by fall. The mean of fall was 1.10 in whole the sample. In terms of neurodegenerative disorder, 2.5, and 1.5 percent of the participants, had dementia and Alzheimer respectively, and less than 2 percent of the sample reported using medicine for neurodegenerative diseases. The mean age was 70.4 with 54.7 percent females, mostly White (60.7 percent) with 44.6 percent high school education.

The regression analysis for the fall analysis showed those with Alzheimer 18 percent were more likely to have fall. Similarly, having dementia was associated with 11 percent higher chance of fall. Males also had 3 percent higher of fall. There was no association between having Alzheimer and injury by fall, however, those with dementia 9 percent more likely to have an injury by fall. Also, Whites were less likely to be injured by fall than other races. Having neurodegenerative diseases were associated with more than 7 times of higher fall numbers. Using Alzheimer perception was a protective factor to reduce the numbers of fall, and higher age was associated with 34% of higher fall numbers.

**Table 1.** Descriptive analysis

|  | Overall<br>(N=8782) |
| --- | --- |
| **FALL** |  |
| No | 7067 (80.5%) |
| Yes | 1715 (19.5%) |
| **Injury by Fall** |  |
| No | 8256 (94.0%) |
| Yes | 526 (6.0%) |
| **Number of Falls** |  |
| Mean (SD) | 1.10 (7.08) |
| **Dementia** |  |
| No | 8565 (97.5%) |
| Yes | 217 (2.5%) |
| **Alzheimer** |  |
| No | 8654 (98.5%) |
| Yes | 128 (1.5%) |
| **Prescription for Alzheimer's Disease** |  |
| No | 8701 (99.1%) |
| Yes | 81 (0.9%) |
| **Prescription for Neurodegenerative Disease** |  |
| No | 8611 (98.1%) |
| Yes | 171 (1.9%) |
| **Age** |  |
| Mean (SD) | 70.4 (10.1) |
| **Gender** |  |
| Female | 4807 (54.7%) |
| Male | 3975 (45.3%) |
| **Race** |  |
| Black | 2282 (26.0%) |
| Other | 1169 (13.3%) |
| White | 5331 (60.7%) |
| **Educational Levels** |  |
| College and more | 3499 (39.8%) |
| High Scholl | 3919 (44.6%) |
| Less than High School | 1364 (15.5%) |

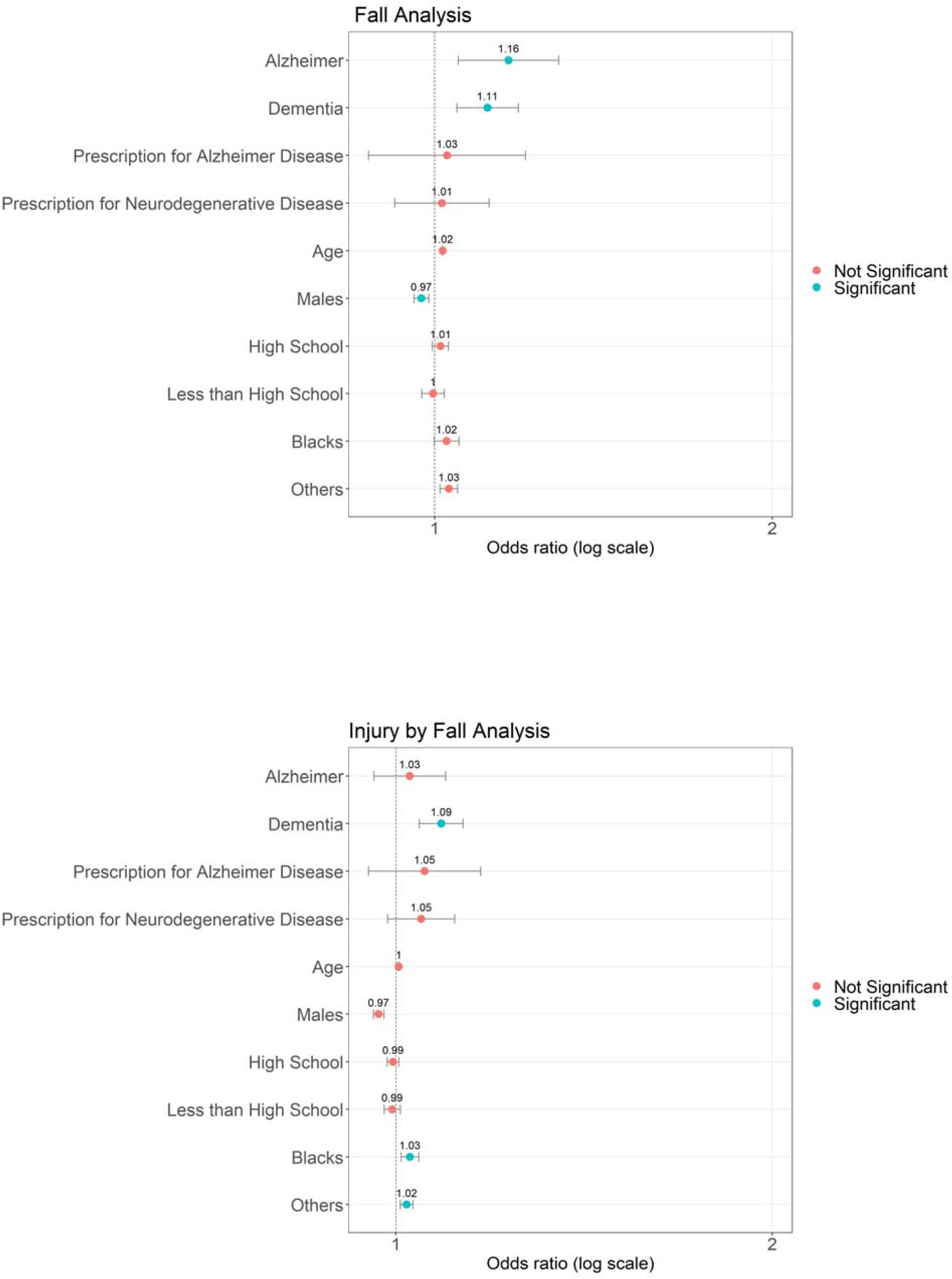
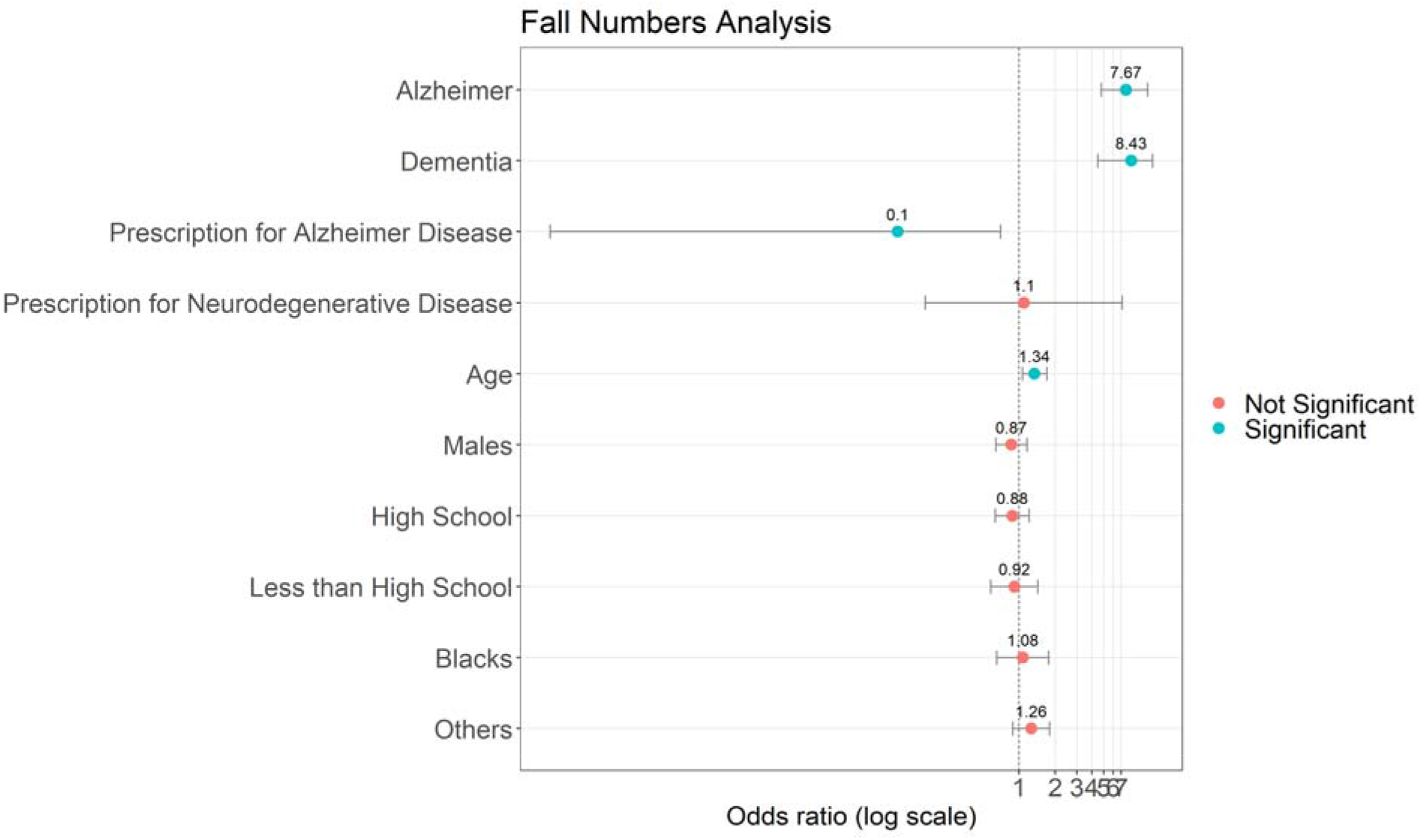

## Discussion

This study aimed to assess the risk of falls among older adults with neurodegenerative diseases and how a prescription for neurodegenerative disease can be a protective factor to reduce fall risk. The result shows older people with dementia and Alzheimer’s were at increased risk of falls and their consequences. In this sample of older people, 20% reported at least one fall, while just a third of them were injured by a fall. Around two percent of the sample had either dementia or Alzheimer, and just one percent of the sample took medicines for neurogenerative diseases. Although this study indicated a more than 10 percent higher risk of at least one fall for elderly people with dementia and Alzheimer, the risk was not higher as in other similar studies20. However, those with dementia and Alzheimer’s had more than 7 times of higher chance of a higher number of falls. In terms of medications, this study showed Alzheimer’s medications were significant for protection to decrease fall numbers.

Among the advantages of this study are the well-characterized sample of older adults as well as information on neurodegenerative medications. This study has the limitation of a homogeneous sample. This sample of older adults was primarily white and well-educated. According to the study, falls are more common among white people compared with minorities and among those with higher educational levels compared with those with lower educational levels.^21,25^ Another possible limitation of the study is the use of self-reported measures of falls. Even though all participants were cognitively normal (CDR 0) and fall ascertainment methods were intensive, underreporting of falls cannot be ruled out. Although strategies were used to aid in recalling fall events, underreporting of falls is more likely with recurrent falls.^22^ Falls were reported at a higher rate than expected in our sample. There is no indication that our enhanced surveillance methods or the characteristics of the sample contributed to an increase in reported falls.

Despite limitations, this study points to the importance of understanding neurodegenerative diseases were associated with higher fall risk, injuries and numbers. It is necessary to conduct additional research to determine the extent of fall risk among healthy older adults to determine the mechanism that causes falls differentially in each group. Continuing research could provide a better understanding of how dementia and Alzheimer’s present clinically and of underlying mechanisms that cause falls.

## Data Availability

All data produced in the present study are available upon reasonable request to the authors
All data produced in the present work are contained in the manuscript
All data produced are available online at
https://hrsdata.isr.umich.edu/data-products/2020-hrs-core

## Notes

### Competing Interest Statement

The authors have declared no competing interest.

### Funding Statement

This study did not receive any funding

